# Performance of and preference for saliva sampling for detection of SARS-COV-2 in the Bahamas

**DOI:** 10.1101/2024.10.10.24314969

**Authors:** Indira Martin, Pearl McMillan

## Abstract

Implementing public health diagnostic modalities that are simultaneously accurate and acceptable is integral to effective pandemic response. In this regard, saliva has proven to be a reliable alternative to nasopharyngeal swabs (NPS) for the detection of SARS-COV-2 infections. In particular, the SalivaDirect protocol utilises untreated saliva as its sample type, and removes the need for RNA extraction, thereby decreasing the time and cost of diagnosis by RT-PCR. IN the current study we piloted SalivaDirect in the context of The Bahamas archipelago, where it demonstrated acceptable performance, with 95.2% concordance with NPS. However, there was discordance in 3 of the 8 total SARS-COV-2 positive samples,all of which were above Ct 30 and therefore presumably of low infectivity. Furthermore, a significant majority of survey respondents chose saliva as their preferred sample type and this was associated with citing ‘discomfort’ of NPS sampling as the reason for their choice. These results support the practical use of SalivaDirect in the Bahamas as a mass testing tool.

## 1. Introduction

Beginning in late 2019, the world has experienced a pandemic caused by the highly contagious respiratory coronavirus SARS-COV-2, necessitating mass testing for the identification and isolation of new cases, in an attempt to interrupt community transmission (1). In this regard, nasopharyngeal swab (NPS) sampling has globally become the ‘gold standard’ method of diagnosis of SARS-COV-2 infection, but this sampling technique often causes discomfort and can cause nasal bleeding. In addition, because it can be technically challenging, it may lead to operator error and potentially false negative results (2-8). There is also substantial cost entailed in sourcing NPS and nucleic acid extraction kits, and reliance on this sampling modality has resulted in bottlenecks to SARS-COV-2 testing in the past-particularly in resource-limited settings (9).

Saliva has been demonstrated as a feasible alternative to NPS sampling for SARS-COV-2 testing, with comparable sensitivity and specificity (10). The SalivaDirect protocol, which is a streamlined, saliva-based RT-PCR methodology that removes the need for RNA extraction, has also reported excellent concurrence with NPS (11). Therefore, in the present study we piloted the Salivadirect SARS-COV-2 RT-PCR testing protocol in the Bahamas-an archipelagic small island developing state in the Caribbean-and surveyed the preference for saliva versus NPS among Royal Bahamas Defense Force military personnel. The overall objective of this study is to assess the performance of SalivaDirect and preference for saliva sampling for routine screening for SARS-COV-2.

## 2. Methods

### 2.1. Study participants

Study participants were visitors to the Royal Bahamas Defense Force (RBDF) sick bay in Nassau, New Providence island, and attendees at a test outreach event on Exuma island, in The Bahamas archipelago. In total, 63 persons were recruited for the study including 29 persons from the Exuma outreach event and 34 from the RBDF. Median age of participants was 33.4 and there were 41 (65%) males and 22 females (35%).

### 2.2. Biological sampling

For each study participant, a NPS and saliva sample was obtained in parallel. NPS were collected according to WHO guidelines (1) and collection of saliva was performed as described in the Salivadirect protocol (11). Briefly, a minimum of 0.2ml of pooled saliva was collected from each participant in a labelled plastic tube and biohazard bag. NPS and saliva samples were transported to the National Reference Lab (Nassau, New Providence Island, The Bahamas) at 4-8C and stored at -70C until RT-PCR testing was conducted.

### 2.3. Survey Questionnaire

A simple anonymous and voluntary survey was designed to assess participant preference for saliva versus NPS, as well as gender and age category. This survey was offered to the nested RBDF sub-group (n=34). The survey is shown in Appendix 1.

### 2.4. SalivaDirect RT-PCR protocol

RT-PCR was performed at The National Reference Lab. Briefly, Salivadirect is an extraction-free protocol in which saliva is mixed with proteinase K and heat-inactivated prior to RT-PCR targeting of the viral N1 gene and the human RP gene. RT-PCR was conducted on the Quantstudio 6 Flex RT-PCR thermocycler. Primer sequences and RT-PCR conditions used are described in ref 11.

### 2.5. NP swab RT-PCR

RT-PCR on NP swabs was performed as previously published (12). Briefly, nucleic acids were extracted from the swabs using the QIAamp Viral RNA kits (Qiagen). RT-PCR was conducted using primers targeting the SARS-COV-2 E gene and the human RNAse P gene on the Quantstudio 6 Flex RT-PCR thermocycler (Thermo Scientific) (12).

### 2.6. Ethics

This study was approved by The Bahamas Ministry of Health National Medical Ethics Committee.

### 2.7. Statistical analysis

Statistical analysis was performed in R statistical software (4.3.3). Fisher’s exact test (two-tailed) was employed to analyse participant preference for NPS versus saliva sampling, as well as associations between independent variables (gender, age and reason for preference) with sample preference.

## 3. Results

### 3.1. Pilot Performance of the SalivaDirect assay

Out of the 63 paired samples, 8 NPS were positive for SARS-COV-2 by RT-PCR and 55 were negative. On the other hand, the Salivadirect protocol resulted in a total of 7 positive test results and 56 negative results. Overall, there were 3 discordant sample results-1 which was positive by Salivadirect and negative by NPS; and 2 that were positive by NP swab and negative by Salivadirect. However, all 3 discordant sample results had RT-PCR cycle thresholds (Ct values) above 30 (Figure 1). 54 samples were SARS-COV-2 negative by both NPS and SalivaDirect. Overall, the qualitative concurrence between the two sampling methodologies was 95.2% (60/63) See Table 1.

**Table 1:** Performance of SalivaDirect compared to NPS RT-PCR.

|  | Salivadirect-Positive | Salivadirect-Negative | Total |
| --- | --- | --- | --- |
| NPS-Positive | 6 | 2 | 8 |
| NPS-Negative | 1 | 54 | 55 |
| **Total** | **7** | **56** | **63** |

**Figure 1:**
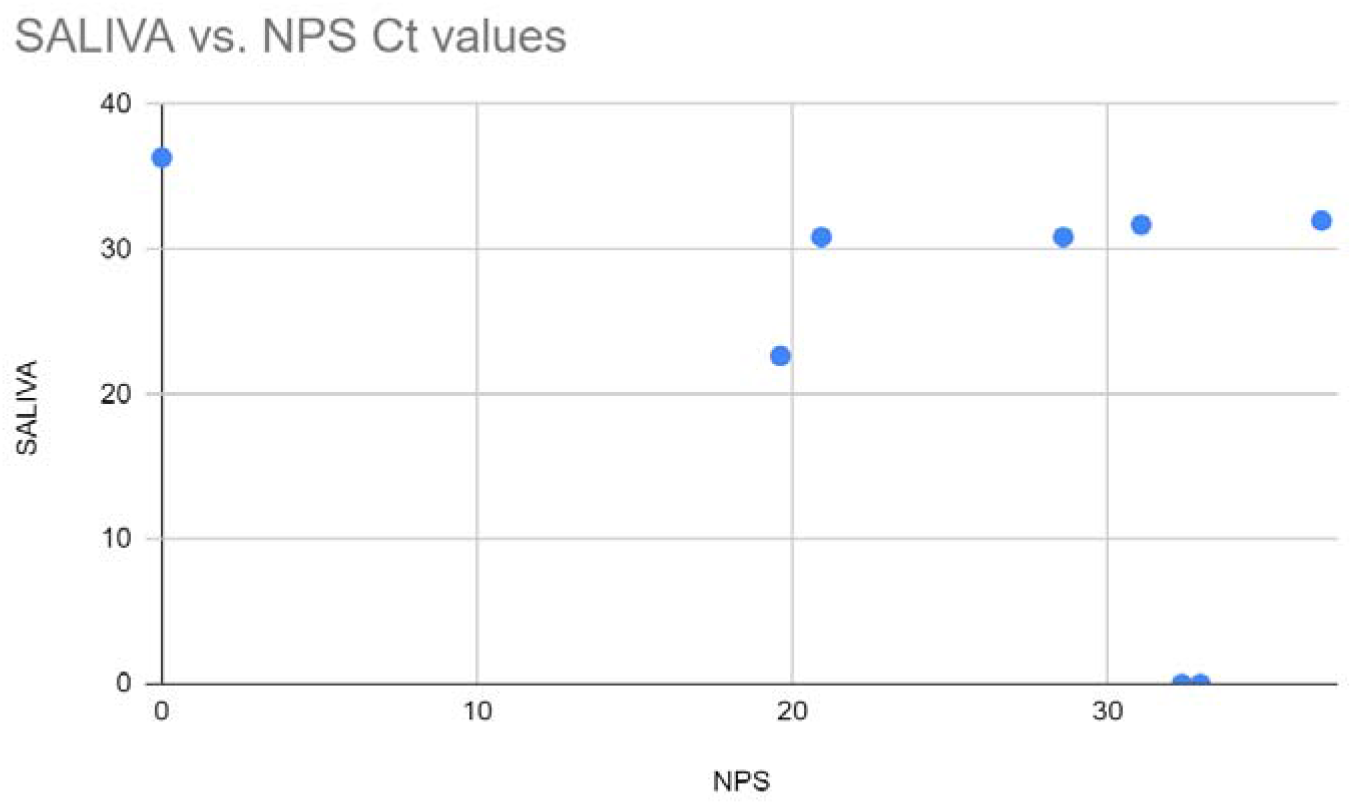
Relationship between Ct values of SARS-COV-2-positive NPS and SalivaDirect samples

### 3.2. Saliva was the preferred sample among survey respondents

Of 34 persons offered the voluntary test preference survey, 27 responded (79%), of whom 24 (89%) were male and 3 (11%) were female. 11 persons (41%) had no preference. Of those who expressed a preference (n=16), a significant majority preferred saliva (87.5%, p=0.004) and this preference was associated with citing less discomfort/pain as the reason for stated preference (p=0.03). See Table 2.

**Table 2:** Preference survey descriptive statistics.

|  |  |  | *p value* |
| --- | --- | --- | --- |
| Total, n | 27 |  |  |
| No preference, n(%) | 11(41) |  |  |
|  | NPS | Saliva |  |
| With preference, n(%) | 2 (12.5) | 14 (87.5) | **0.004** |
| Age |  |  |  |
| <40 | 2 | 6 |  |
| >40 | 0 | 8 |  |
| Gender |  |  |  |
| Male | 2 | 12 |  |
| Female | 0 | 2 |  |
| Reason |  |  |  |
| Faster/more efficient | 1 | 1 |  |
| Accuracy | 1 | 0 |  |
| Less discomfort | 0 | 12 | **0.03** |

## 4. Discussion

Implementation of diagnostic modalities that are both sufficiently accurate and acceptable to potential target populations is integral to any public health response to infectious disease threats. In this regard, saliva has emerged as a key diagnostic tool for a broad range of diseases (13), that may be particularly useful for reaching geographically isolated, hard to reach and/or resistant populations for testing and re-testing. This is because it does not require specialised collection devices, and is stable for long periods at room temperature or higher (11,14). In the case of SARS-COV-2, as a sample type saliva yields comparable results to NPS (8-11), and is therefore a key but under-utilised biomedical tool for detecting SARS-COV-2-and potentially other respiratory viruses (15).

Of course, like any sample type, saliva has its potential technical drawbacks which must also be considered. For example, it has been reported that there is a possible matrix effect of saliva that has not undergone RNA extraction as is the case with the SalivaDirect protocol we piloted in the present study-ie that it may contain PCR inhibitors (16). Another consideration is viral tropism in saliva versus the nasopharynx, and apparently this can differ according to which SARS-COV-2 variant is present (17,18). Indeed, in our study sample, we saw evidence of this effect, since we detected SARS-COV-2 in two NPS samples that were negative by SalivaDirect, and in one SalivaDirect sample that was negative by NPS (Table 1, Figure 1).

Nevertheless, the overall concurrence between SalivaDirect and NPS was high at 95.2% (ie 60 of the 63 paired samples received the same qualitative RT-PCR result), and the three discordant samples were all above Ct 30 (Figure 1), indicating that these samples were of low infectivity and presumably low clinical relevance, given that samples of Ct values greater than 30 are not cultivable in vitro (19-22). We found that there was an overall concurrence of 95.2%, with a positive concurrence of 62.5% (5/8 samples) and a negative concurrence of 96% (54/56 samples), which was consistent with previous reports of the performance of saliva as a diagnostic sample for SARS-COV-2 detection (8-11), and more specifically with the SalivaDirect protocol (11,23-25).

Notwithstanding the satisfactory performance of saliva for SARS-COV-2 diagnosis, effective public health diagnostic interventions are necessarily successful in both the biological and behavioural domains, meaning that they must be both accurate and acceptable to the target population. We assessed the latter parameter in military personnel-whom are at high risk due to their frontline service and wide distribution throughout the Bahamas archipelago in times of emergency. Although 41% of respondents to the preference survey had no preference between NPS and saliva, a significant majority (14 of 16) of those with a preference selected saliva as the more acceptable sample type. Additionally, a preference for saliva was significantly associated with citing ‘less discomfort’ as a reason for the selected preference, demonstrating that comfort was a significant consideration among people who preferred saliva sampling (Table 2). Therefore these results in our context are consistent with those seen elsewhere, indicating that saliva may be a feasible sample type that is preferred by many persons within target groups, and may therefore increase the probability of testing and re-testing (26-32).

Our study was limited by a small sample size and a predominance of SARS-COV-2 negative participants. Nevertheless, we captured some of the nuance associated with the performance of saliva-based testing in the unique context of the Bahamas. We show that the RT-PCR results seen in our SalivaDirect pilot were comparable with those obtained from testing NPS, and saliva was the preferred sampling modality among military personnel who expressed a preference. This supports the use of the SalivaDirect protocol and argues for the wider utilisation of saliva in diagnosis and surveillance in the covid-19 pandemic, also warranting its consideration in future public health emergencies.

## Data Availability

All data produced in the present study are available upon reasonable request to the authors

